# “What witchcraft is this?”: Paramedics report gains in productivity, well-being, and patient flow from piloting ambient voice technology in an NHS Ambulance Service

**DOI:** 10.64898/2025.11.27.25340564

**Authors:** David Davis, Jo Wray, Geralyn Oldham, Daniel Elkeles, Timothy Borham, Paul Gough, Maya Tomlinson, Nick James, Alastair Hill, Eleanor Sullivan, Stephen Mathew, Robert Robinson, Jasmine Balloch, Dom Pimenta, Sarah Kirk, Anna Booth, Andrew M. Taylor, Shankar Sridharan

## Abstract

**Objective:** UK ambulance services face record demand, resourcing challenges and rising clinical documentation burden. Ambient voice technology (AVT) coupled with generative AI offers a potential solution by automatically transcribing consultations and drafting clinical notes. Our aim was to test AVT in a complex and undifferentiated pre-hospital context.

**Design:** Within-subject pre-post evaluation

**Setting:** Two operational pathways (remote telephone triage in the Clinical Hub (“Hear and Treat”) and on-scene care provided by paramedics (“Face-to-Face”) in a large metropolitan ambulance trust.

**Participants:** Clinicians in the Clinical Hub and paramedics providing on-scene care

**Intervention:** Ambient voice technology

**Main outcome measures:** Assessment duration; patients handled per hour or shift; documentation quality; on-scene, handover and combined incident ‘job cycle’ times; clinician cognitive load (NASA-TX).

**Results:** 656 Hear and Treat cases were managed with AVT. Compared with baseline, AVT shortened assessments by two minutes (15 vs 17 minutes) and increased throughput by 15% (2.3 vs 2.0 patients/hour) without compromising documentation quality. In Face-to-Face care (n=344 AVT contacts) mean on-scene time decreased from 44 to 41 minutes (-6.8%), while median improvement was 15.2% (46 to 39 minutes). Combined on-scene + handover-to-clear time reduced by 2.8 minutes (-4.8%). Across both settings there was a significant reduction in clinician cognitive load, improvement in clinician experience and perceived benefits to patients.

**Conclusion:** This evaluation demonstrated an increase in remote telephone triage throughput and a reduction in on-scene care times with AVT. Scaling such improvements would translate into real-world operational gains that, if implemented nationally, would ease pressures on ambulance services whilst improving patient care. AVT demonstrably frees clinical time across remote ambulance workflows, aligning with national ambitions to “bulldoze bureaucracy” and speed-up care. Strategic scaleup with robust governance could release capacity equivalent to thousands of additional patient contacts each day and improve outcomes for some of the most unwell patients.

## Introduction

Ambulance services sit at the sharp end of NHS pressures: 999 calls have doubled in a decade while response-time targets are routinely breached.^1–5^ In April 2025, the Department of Health & Social Care (DHSC) hailed AI assistants as a “game-changer” for reducing administrative tasks and seeing more patients in A&E and beyond.^6^ Parallel multi-site NHS evaluations have shown AVT increases direct care time by 23.5□% and cuts emergency-department documentation time by 51.7□%.^7,8^

Less is known about the application of AI in ambulance workflows, where documentation must often be completed under time pressure, in noisy environments and frequently after the patient hand-over. Ambulance clinical practice and context are highly varied, largely undifferentiated and presents a range of complexities compared with practice in other clinical settings. From elderly fallers and non-transport decision making to high acuity situations like cardiac arrest and acute behavioural disturbance, ambulance clinicians regularly face symptoms that do not fit standard categories and instead have to make sometimes complex and ambiguous decisions. Clinical consultations take place within highly varied environments, from people’s homes, to the scene of busy incidents, such as a train station, night club or side of a motorway – where AVT has not previously been tested.^9–11^ Furthermore, unlike the hospital environment, paramedics are responsible for the care, assessment, treatment, medicines administration and clerking, often whilst a colleague drives – the workflow is like no other healthcare workflow. Ambulance leaders therefore describe AI adoption as “life or death” and emphasise the importance of “getting it right first time”.^12^

The ambulance and pre-hospital workforce is also known to be highly diverse and has seen over the last twenty years huge professional advancement and social mobilisation. Whilst comparators are hard to identify, the sector is known to employ a large number of neurodiverse individuals and, much like other sectors, has part of its workforce which is older and may find adoption of digital technologies challenging.^13^ ^14^

Our aim was to evaluate use of AVT in both remote telephone triage (Hear and Treat) and frontline ambulance care (Face-to-Face) within a large NHS ambulance trust in England, as part of a wider study trialling AVT across multiple health-care settings.

## Methods

### The AVT technology

Great Ormond Street Hospital (GOSH), who led and coordinated the wider trial across all sites, entered into a partnership in 2023 with Tortus AI, a UK health-tech start-up company, to transcribe consultations and auto-generate clinician-specified notes. Tortus AI was selected for its single-supplier consistency, rapid frontline responsiveness, deep understanding of NHS data-protection, cybersecurity, clinical safety and model-governance requirements, and completion of independent certifications (CE Plus, bias assessments, medical-device accreditation). Information governance approvals were obtained.

### User Selection

An open call for expressions of interest was circulated across the clinical workforce, using the internal intranet, to recruit participants for the pilot; there was no cap on participation numbers. Clinicians who expressed interest were provided access once they completed the accompanying pre-use survey. Upon enrolment, users received access to their AVT account credentials as well as training materials to begin using the AVT tool.

**Design and settings** – A within-subject pre-post evaluation was conducted across two operational pathways:

1. **Hear and Treat** – clinicians (paramedics and nurses) in the Clinical Hub (CHUB) manage a broad range of patients by telephone. Two AVT-enabled workstations were installed.
2. **Face-to-Face** – paramedics and non-regulated ambulance clinicians in double-crew ambulances and fast-response units trialled a mobile AVT application during routine emergency calls.

## 1. Hear and Treat

### Implementation

To enable AVT use in the CHUB setting, additional hardware was required to route telephone audio into the AI system. For the pilot phase, two workstations were equipped with this setup—limiting concurrent use to two clinicians out of approximately 50 available workstations and an average workforce on duty of 28. This limitation was implemented both as a safety measure and for technical reasons.

Having approved, at an organisational level, a template for documentation, pilot users were asked to carry out their shifts as they would normally, incorporating the AVT to generate documentation, validated for accuracy by the clinicians, for each call they assessed; the only exclusions were calls where the patient did not consent to AVT use.

CHUB practice involves clinicians reviewing an assigned call queue, organised by ambulance service operational sector. These calls are reviewed by a clinical manager using the available information from the 999call for their suitability for a remote assessment. The calls marked as suitable are reviewed in order by the next available clinician based on time in queue and clinical presentation. No restrictions were imposed on case selection during the pilot, the AVT tool was used across a full spectrum of cases presenting in routine practice, providing consent was obtained. AVT created templated driven standardised records which were copied into the electronic record system following clinician validation.

### Data Collection and Cohort

Data were collected during April 2025 using pre-existing CHUB performance monitoring tools and data reports. Metrics included:

- Sample size
- Assessment duration
- Assessments per hour
- Documentation quality

## 2. Face-to-Face (Ambulance-Based Assessments) Implementation

Users accessed the AVT technology on existing tablets already used for clinical documentation and other digital clinical applications. A standardised template was designed and internally approved for use. Users were allowed to add to these templates using the included no-code designer within the AVT software, but not to remove any key headings that had been included. Use of AVT was either in an ambient listening mode of the clinical assessment and consultation or a post-incident dictation. Users generated the templated notes and, following clinician validation of the content, copied the entire record into a free text note and the relevant context into semi-structured fields within the electronic record. The AVT technology was primarily used in ambient consultation, however, clinicians were at liberty to use post-episode dictation after the episode of care had completed.

### Data Collection and Cohort

A six-week post-implementation evaluation (5/3/35-14/4/25) was compared against a three-month pre-implementation baseline (2/12/24-2/3/25).

Metrics were:

- **On-scene time** (for conveyed patients; period spent with a patient prior to transport)
- **Handover-to-clear time** (hospital handover to vehicle availability)
- **Combined time** (on-scene + handover-to-green (green: ready for dispatch to next patient)
- **See and Treat on-scene time** (for non-conveyed patients (total time with patient when they are not conveyed to another place for treatment)

Of note, across the whole organisation a one-minute improvement in average on-scene time (for conveyances) from pre to post implementation was observed, likely due to service improvement activities. To avoid incorrectly attributing reductions in time to the AVT, one minute of clinicians’ on-scene time improvements was attributed to wider improvement within the trust and not the technology.

### Evaluation of clinician experience

#### Survey data

Clinician experience measures were designed for both baseline and AVT and included *‘what three words’* to capture their experience of their most recent shift, Likert-scale questions on experience, confidence, computer tasks, time with patients, and documentation quality together with a Net Promoter Score.^15^ Internal reliability was good (Cronbach alpha =.856). The NASA Task Load Index (TLX) was also included.^16^

#### Interview data

Clinicians were invited to participate in online individual interviews to share their experiences of AVT. Discussions were informed by a topic guide, audio-recorded with consent and transcribed verbatim.

### Analysis

Time and patient number data and clinician experience measures were analysed using descriptive statistics. Baseline and AVT survey scores and scores on NASA TLX were compared using Wilcoxon signed-rank tests. A total TLX score was computed by summing the subscales without weighting and dividing by the number of subscales (Raw-TLX)^17^. A p value of <0.05 was considered statistically significant. Interview data were analysed using the Framework approach^18^.

### Study approval and consent

The overall study was registered at GOSH as a clinical evaluation study and local approvement and study registration were additionally undertaken. NHS ethical approval was not required. Participating clinicians provided verbal consent; consent for surveys was assumed if a completed survey was submitted. Patients in the ‘Hear and Treat’ pathway were informed that calls were being recorded and transcribed and that they could ask for transcription to be deactivated. Use of AVT in the face-to-face arm, where patients were present, was subject to verbal consent from patients and others on scene, including ambulance staff not in the trial. Some situations meant that clinicians were permitted to use discretion for AVT use depending on clinical circumstances, patient presentation (including mental capacity), or environmental factors.

### Patient and public involvement

There was no patient/public involvement in the work reported here.

## Results

### Hear and Treat (Clinical Hub)

There were 11 pilot users. Due to variation in shift patterns, some days had no AVT users on duty; at other times, demand exceeded the number of equipped workstations. AVT users completed 656 assessments (2.7% of total CHUB activity), while non-users completed 23,044 assessments. No exclusions were applied to the samples.

***Duration of patient assessments:*** AVT supported assessments averaged 15 minutes, compared to 17 minutes without the tool, resulting in a 2-minute (11.8%) improvement (Figure 1).

**Figure 1:**
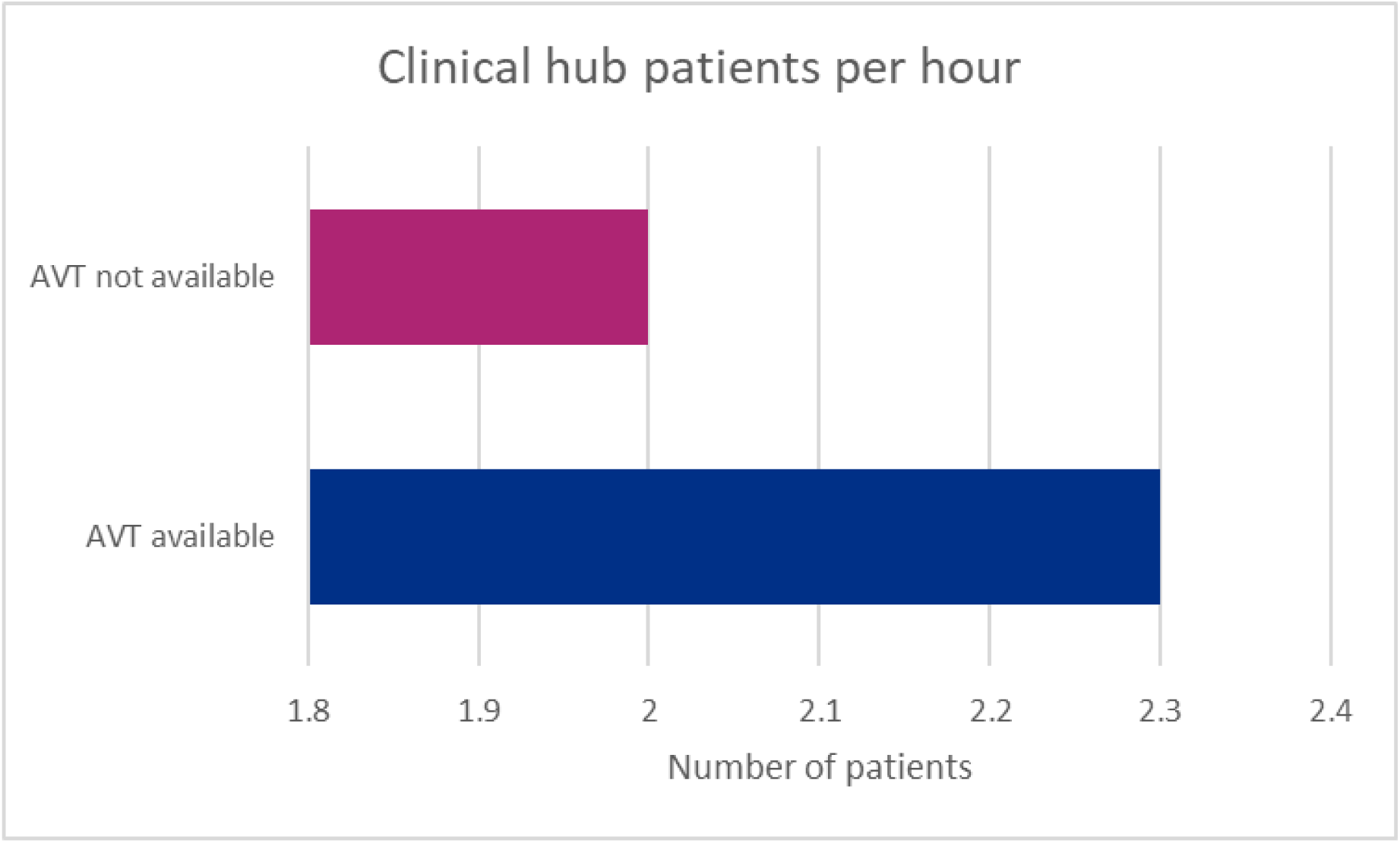
Number of patient assessments completed by the Clinical Hub with and without AVT.

**Productivity:** AVT users assessed an average of 2.3 patients per hour compared to 2.0 among non-users, a 15% increase in the productivity of the AVT-enabled clinicians.

**Documentation Quality:** CHUB documentation quality was evaluated using existing business-as-usual audit processes (supplementary material Box A). To provide assurance around the safe implementation of the AVT tool, additional audits were carried out during the first two weeks of the pilot using the existing call audit process.

Audits demonstrated consistent documentation quality across both groups, with AVT users showing a slight improvement in average audit scores: 98.5% vs 98.2% for non-users.

### Face-to-Face (Ambulance-Based Assessments)

The pre-implementation cohort comprised 816 patient contacts; the post-implementation cohort included 344 AVT-documented cases from seven clinicians.

## Primary Outcomes

- **On-scene time (mean):** Decreased from 44 to 41 minutes (6.8% improvement). Individual clinician improvements ranged from a 1-minute increase to an 8-minute reduction.
- **On-scene time (median):** Reduced from 46 minutes (IQR 33–54) to 39 minutes (IQR 35–45) (15.2% improvement).
- **Handover-to-clear time (mean):** Increased from 14 to 15 minutes.
- **Combined time (mean):** Reduced from 58.4 to 55.6 minutes (4.9% time saving).
- **Combined time (median):** Reduced from 58 to 54 minutes (6.9% improvement).

These data demonstrated net productivity gain for the clinicians after AVT implementation.

### See and Treat Subgroup

In non-conveyed cases, on-scene time improved by an average of 4 minutes (3.8%). In one instance, a clinician reduced average on-scene time by over 22 minutes, though this could reflect patient variation rather than a technology effect or some other variable.

### Patients per Shift

- **Mean:** Largely unchanged (0.7% decrease).
- **Median:** Increased by 0.2 patients (4.2%), from 4.8 to 5.

### Solo Responder Vehicles

An additional analysis was conducted of five clinicians who were assigned to Solo Response Vehicles, which are non-conveying assets focused on rapid on-scene assessment and treatment. The evaluation demonstrated improvements in clinical productivity when using AVT across measures of patient throughput.

Key findings:

- **Patients per shift (mean):** Increased from 4.25 to 4.5 (5.9% improvement).
- **Patients per shift (median):** Increased from 4 to 5 (25% gain).
- **Patients per hour (mean):** Increased from 0.375 to 0.45 (20% rise).
- **Patients per hour (median):** Increased from 0.35 to 0.45 (28.6% gain).

### Survey findings

Twenty-nine clinicians across both pathways completed surveys at both baseline and with AVT, the majority (n=18; 69%) of whom had used AVT before and strongly agreed/agreed that they felt confident to use it (n=28; 97%). There were significant positive changes in the level of agreement that they had sufficient time with each patient (Z=-2.179; p=.029), were able to give patients their full attention (Z=-3.262; p<.001), satisfaction with care given (Z=-2.951; p=.003) and their overall experience was positive (Z=-2.514;p=.012) in the AVT compared with baseline condition (Figure 2(a)).

**Figure 2:**
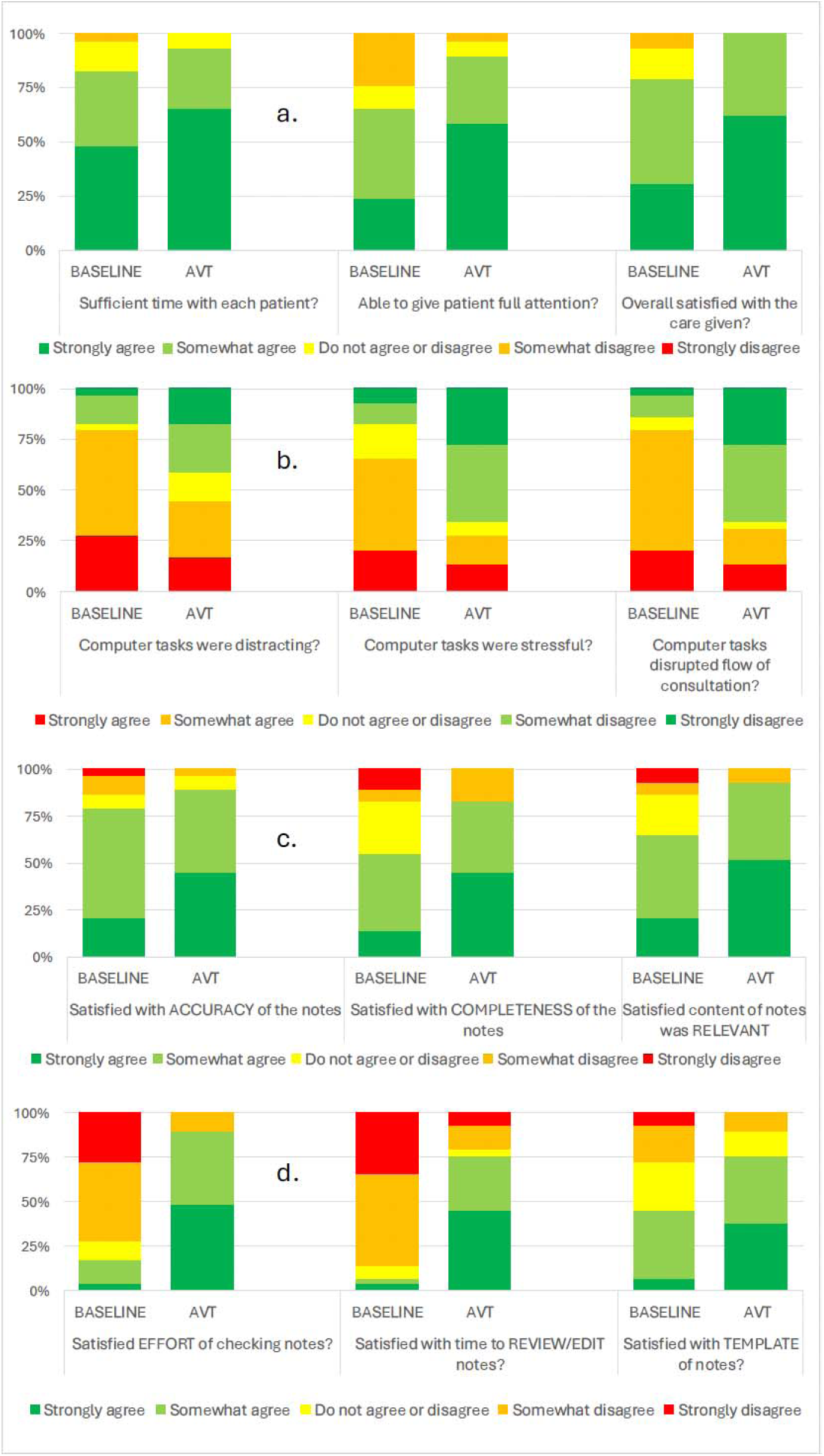
Clinician experience: aspects of care given (a), computer tasks (b) and elements of the clinical note made during patient episodes (c and d): baseline vs AVT.

For questions about using the computer during clinical encounters, responses were more positive for AVT (distracting: Z=-2.583; p=.010; stressful: Z=-3.153; p=.002; disrupted flow (Z=-3.511; p<.001) (Figure 2(b)). All aspects of the clinical notes were rated more positively with AVT (satisfaction with accuracy: Z=-1.986; p=.047; completeness: Z=-2.464; p=.014; relevance: Z=-2.826; p=.005; effort of checking the notes (Z=-4.224; p<.001), time to review/edit notes (Z=-4.051;p<.001) and notes template (Z=-3.036; p=.002) (Figure 2c and d).

A higher percentage of patient notes was completed at the end of the encounter with AVT (Figure 3).

**Figure 3:**
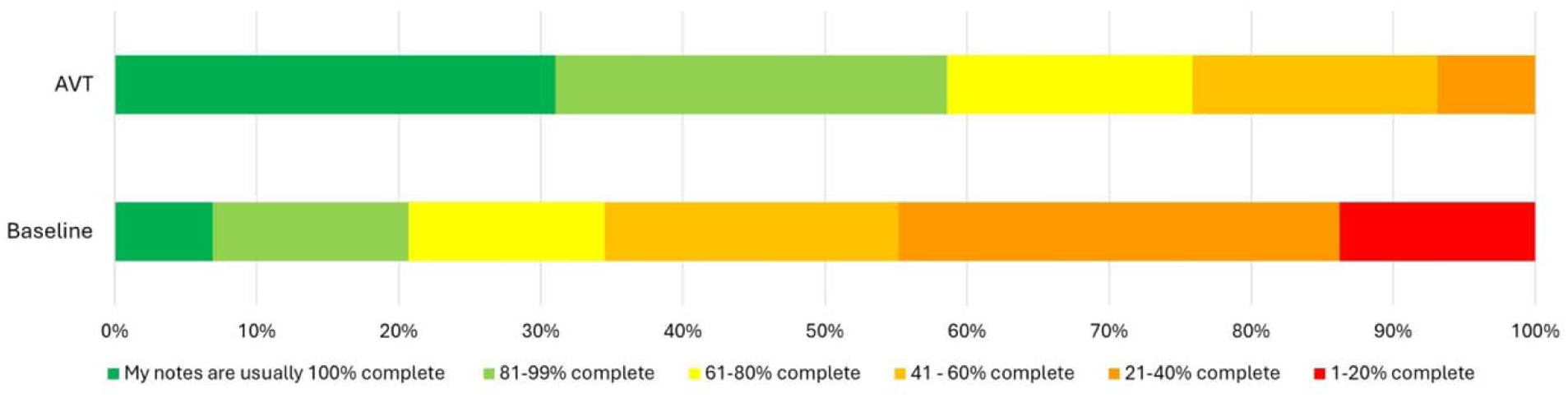
Percentage of notes completed at baseline and with AVT by the end of the encounter.

“*What three words”* are shown in Supplementary Material.

### NASA Task Load Index

There was a significant reduction in total NASA score with AVT (baseline median: 5.83; IQR: 1.50; AVT median: 4.42; IQR: 2.21; Z=-3.556; p<.001), with significant reductions in 4 of the 6 subscales – physical demand (Z=1.953; p=.051) and performance (z=1.743; p=.081) were not significantly different (Figure 4).

**Figure 4:**
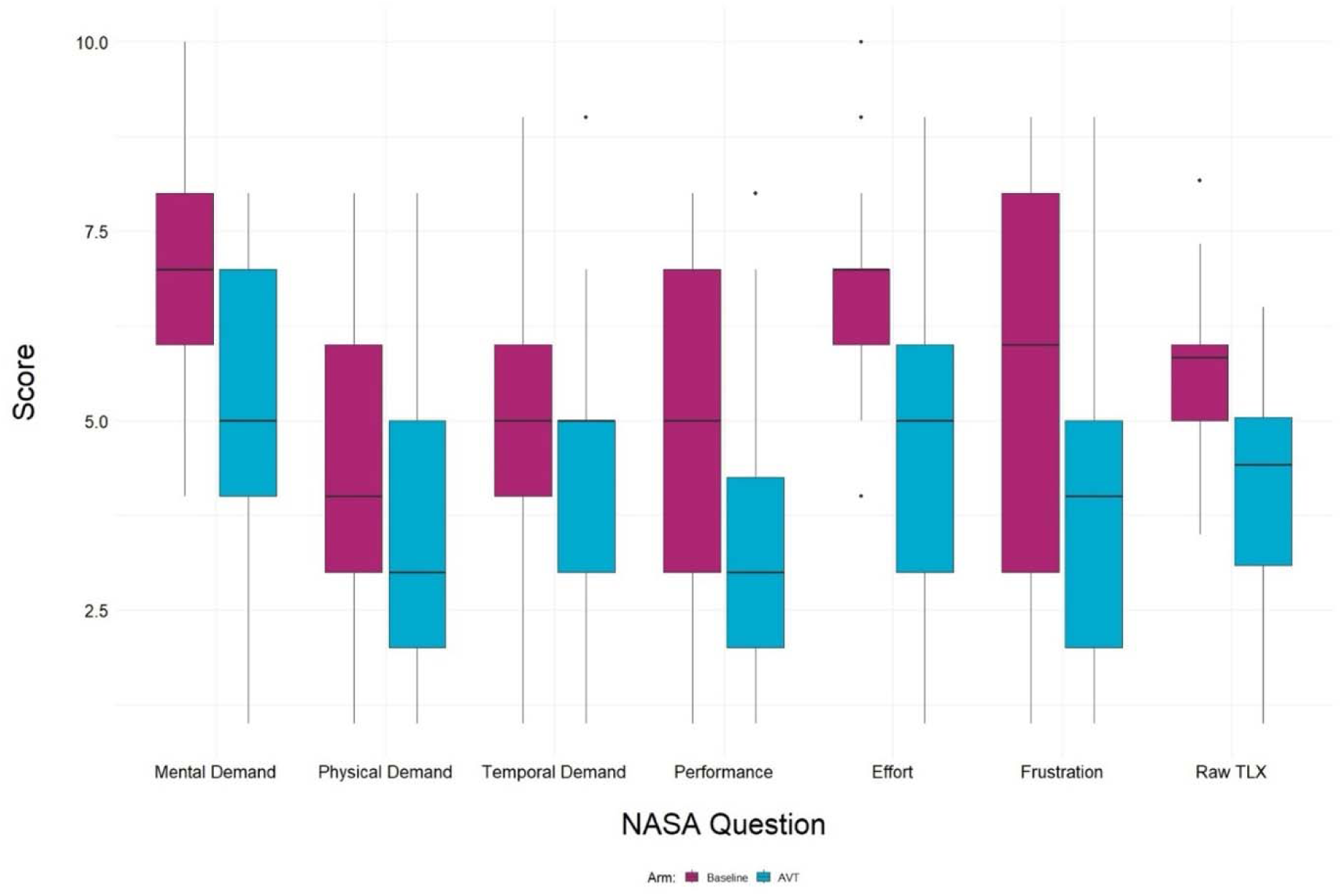
The NASA Task Load Index (NSA-TLX) at baseline and AVT (n=29)

### Interviews

Seven clinicians participated in individual online interviews. Four inter-connected themes and ten subthemes were identified (Figure 5).

**Figure 5:**
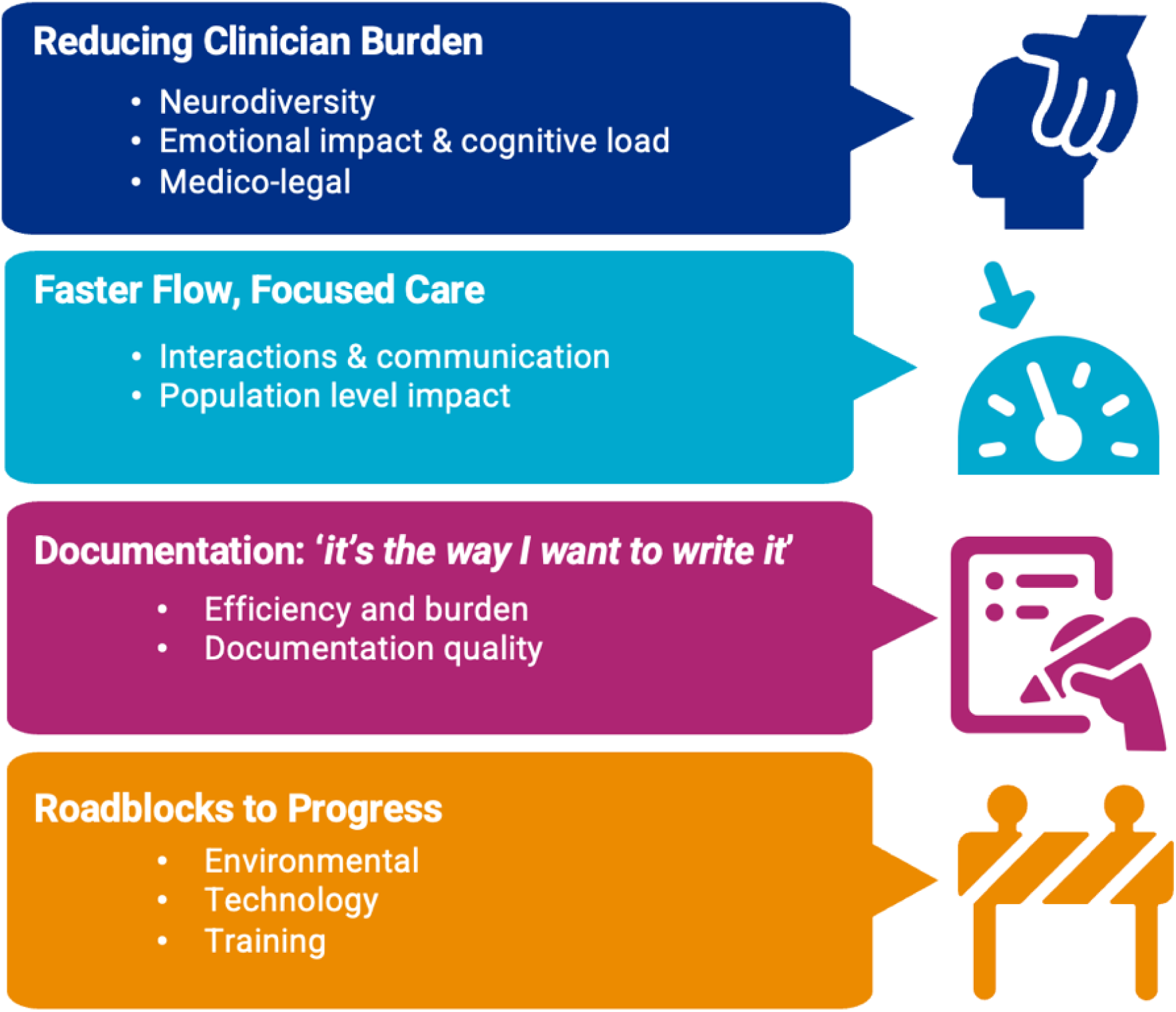
Themes and subthemes from the seven clinician interviews Brief descriptions with illustrative quotes are provided in Table 1.

**Table 1:** Themes, subthemes and participant quotes.

| Theme | Subtheme | Participant quote |
| --- | --- | --- |
| Reducing clinician burden | *Neurodiversity*<br>AVT was seen as particularly beneficial for neurodivergent staff; they described AVT as transformational, allowing them to articulate their clinical reasoning more clearly and with less anxiety; reduced cognitive load, enhanced | “[Before AVT] I would have had to have remembered everything. I suffer from dyslexia and it’s audible dyslexia so I’ve had to learn some techniques as to remember stuff for handovers and I do it in a set way...then I’d have to go away and type it at the end of it and remember all of that stuff – it was |
|  | clinical documentation and better patient interaction were all seen as supporting neurodivergent clinicians and their ways of working and some also described benefits for their own self-esteem of having better paperwork. The reduction in typing and paperwork when with a patient was particularly helpful for clinicians with ADHD, who might find it difficult to split their focus between interacting with the patient and writing in the notes. | difficult... with AVT it is so much easier" (C1)<br>"This makes doing my notes...enjoyable rather than a slog. It makes a big difference to me because I could be going over this, giving me a headache, making sure my notes are right..." (C6) |
|  | *Emotional impact and cognitive load*<br>Clinicians described how AVT helped reduce the mental strain associated with traditional documentation processes and the need to remember all details from a consultation. They talked about the high cognitive burden paramedics face during patient care, due to the fast-paced nature of the work, with resulting stress to clinicians – AVT reduced this. Although clinicians mainly discussed reduction in cognitive load as a result of reduced documentation time, for some cases they found it more cognitively intense, which some suggested was due to patient complexity and the time required to validate or refine AI-generated notes. | "It's been really good because it hears things that I've completely forgotten. I think that's one of the best things about it" (C7)<br>"I feel like I am more rested at the end of a shift, less stressed. I know that it's all captured as well" (C5) |
|  | *Medico-legal*<br>Clinicians talked about the value of AVT in improving accuracy, reducing | "Medico-legal is nearly always about what you've recorded not what you've done...the better you get the records |
|  | 'short-cuts' and errors (e.g. through reducing clinicians' copying and pasting from other documentation) and the potential benefit for court cases or if patients/other clinicians question what happened during an encounter. | the less you see of this [legal action]" (C2)<br>"...an objective third party that can evidence that that's what happened, which helps to provide some clarification if, 'no they didn't do an ECG on me' or 'no they didn't listen to my chest' but then AVT has presented that that did happen" (C4) |
| Faster flow, focused care | *Interactions and communication*<br>Clinicians noted that by reducing the need to focus on typing, AVT helped them engage more with patients and family members and that verbal articulation for the AVT recording also fostered inclusive communication and helped to align patients' and families' expectations. This was particularly helpful for patients with mental health issues. They also commented about improvements in their history taking so that the AVT picked it up and that the AVT remembering something they had forgotten – e.g. about a patient struggling with their medication – might prompt further action on their part such as a call to the pharmacy, with potential benefit to the patient. | "You're not looking at your screen and typing – you're just there talking to the patient" (C5)<br>"It helps keep patients and relatives informed... we usually talk in jargon but now I say everything out loud" (C3)<br>"The more accurate clinical record you get, the more concise clinical record you get, the easier and smoother it is for transfer. So you see less error, you see less risk if the clinical record is better (C2)<br>"The one time that I didn't have it [AVT] I was typing...it was a mental health patient and the lady was a little bit standoffish that I was not talking to her throughout" (C1) |
|  | *Population level impact*<br>Some clinicians also reflected on benefits at more of a population level in that other patients would benefit from improved efficiency with the current patient. One of the CHUB clinicians described how their reduced fatigue and cognitive load as a result of the AVT increased the number of calls they were taking, with benefits for those waiting to speak to someone who was therefore responded to more quickly. | “If I’m seeing the next patient quicker or I’m able to see another patient, there’s an obvious clinical benefit” (C3)<br>“I would say it’s probably upped my calls by about 30%, not only because I’ve cut down on the admin time but mostly because by not having the fatigue from having to write and do everything else, you’ve then got the ability to be able to go on and have more patient interactions from doing it’ (C5)<br>“If you take 10 minutes off a job cycle and you do that five times a day, that’s another patient you’d be seeing that day. That’s a patient that’s getting an ambulance response quicker...if you reduce your overall response time to a category one cardiac arrest patient by one minute, you see an extra ... chance of survival” (C2) |
| Documentation:<br>‘It’s the way I want to write it’ | *Efficiency and burden*<br>Clinicians consistently reported that AVT significantly reduced time and effort required to complete clinical documentation; verbalising rather than typing notes allowed more efficient transitions between patients; this was often connected to reduced job cycle times and the potential to attend more patients in a shift, although this was variable. Reduction in documentation time was also seen not only as a | “It generally takes five minutes for me. Five minutes of verbalising my notes. Before, it could easily be 45 minutes” (C6)<br>“When I got back in the car and I pressed generate documents, beyond unbelievable” (C3)<br>“Why are we asking our colleagues to have their chill time or their downtime when they’re writing paperwork? No, |
|  | productivity gain but as an opportunity to reallocate time so that clinicians had a break, providing an opportunity to redesign staff workflows around their wellbeing. Some concerns were raised about potential future pressures to reduce job-cycle times due to improved AVT-driven efficiencies and unrealistic expectations from management. | we need to be better. If we can say, look, we're so much better at doing paperwork now because of [AVT] we can give you another break" (C3) |
|  | *Documentation quality*<br>Clinicians described paperwork as being more coherent and grammatically correct with AVT but some instances were described of hallucinations or mis-hearing, particularly in more chaotic settings. Although some talked about over-summarising, others described how the summarising did differentiate between clinically relevant and irrelevant social information, for example. Template set up was not always providing clinicians with what they wanted although some described their own workarounds, to good effect. A 'one size fits all' template was not seen as a good idea because of variations in practice and loss of the individual clinician voice. Linked to documentation accuracy was the recognition of the importance of clinicians reading and checking the output. | "I just dread it [paperwork] because it seems to take me so long to word things. And that's actually one thing that I love with the AI, is the way it words things in such a great way. It sounds really good and it's not the way I would have thought to write it but it's the way I want to write it" (C7)<br>"I'm not convinced the template modelling is quite where it needs to be... I can't quite get it to template exactly how I want it to... People need to realise they need to read it. It's still their record. It may have a mistake in it – AI is not foolproof" (C2)<br>"I've set up my own template and it actually does a good job of filtering out most of the stuff. Obviously we have a joke and a laugh with the patients, it doesn't put that in as well" (C1) |
| Roadblocks to progress | *Environmental*<br>The AVT did not work well in all settings for all clinicians, particularly environments that were more chaotic and when a patient was a higher acuity patient, although others reported that the AVT did work well in such situations which was attributed to a dedicated microphone and user awareness of how to communicate in that setting so that the AVT picked it up. Clinicians also discussed the challenges with other people being present and the AVT incorrectly assigning something a relative said about themselves as being about the patient or the AVT being confused by multiple perspectives. | “.. with a low acuity or moderate acuity patient, in a relatively quiet environment, you run it in AI mode, and you do the ambient listening and that works well. The problem is you can't do that with an ultra-high acuity patient. And you can't do that in a noisy setting. Actually, what we have discovered is it's pretty good with background noise but once you get into a street it's not the same – it just doesn't really work in that setting. But then you move back to that model of ‘take the history and dictate to it’” (C2)<br>“I used it on a major trauma... with police, with multiple agencies and multiple people talking and it captured everything” (C1) |
|  | *Technology*<br>There were a few instances of the software crashing or hardware issues, particularly with microphones. The need for integration with electronic patient records was frequently mentioned. Clinicians also described frustration with not being able to edit the document once it had been finalised. | “Last week there was one where it just completely crashed and didn't load the paperwork and I lost it. But that's only a couple of occasions that's happened...on the whole I would say it's pretty, pretty robust” (C7)<br>“Once you've finalised it under consultation to create the document you can't edit it, you can't rechange it... you might do it and think ‘I've missed out that whole conversation’ and you've got to think ‘well I don't want to rewrite everything. I don't want to re-verbalise everything I've just done” (C6) |
|  | *Training*<br>The importance of sufficient training was highlighted – some clinicians received minimal or no formal training, making it more challenging for them to understand and customise the templates to meet their needs. A number of staff had developed their own workarounds with the templates, such as including more context, and a mechanism for sharing those would have been beneficial. | “There was a meeting but it was first come, first served. I didn’t get on it” (C7) |

Clinicians generally had a very positive attitude to AVT and were keen to describe its potential to colleagues – as one clinician said, “*I said to people, just go and use it…I said to her, just text me when you’ve used it and she sent me a GIF which says, ‘What witchcraft is this?’ And that was her [AVT] review. So, I said did you enjoy using it? Gonna be useful? She said, very much so. It’s very clever.*” As another clinician summarised, *“I genuinely don’t think I would be happy losing AVT. I’m still in control as a clinician. I’m still treating people. It’s just helping me with the paperwork side of it…it means that I’m a lot less stressed*”.

## Discussion

### Statement of principal findings

The evaluation of AVT in an NHS Ambulance Service demonstrated a 15% increase in remote patient triage throughput and a 6–15% reduction in on-scene care times. These improvements reflect the potential for real-world operational gains that could be scaled nationally to improve patient care, ease pressures on ambulance services and emergency departments alike. If a larger scale pilot demonstrates the same or even improved benefits, this is likely to have a positive impact on patient outcomes and care (already suggested in the clinician interviews) and a positive impact on clinicians in their practice, particularly those who are neurodivergent.

In the CHUB, clinicians using AVT completed telephone assessments more quickly, likely as a result of reduced time documenting during the consultation, and managed a greater number of cases per hour. These findings suggest that, even when using interim technical solutions and without full integration of the technology into existing systems, AVT can safely support clinicians working in high-throughput, time-sensitive environments. In the face-to-face ambulance setting, modest but consistent reductions were observed in on-scene and combined clinical times. These improvements were particularly notable in non-conveying units, such as Solo Response Vehicles, where the flexible, mobile nature of the technology aligned well with the clinical context. Whilst the scalability of the data is limited by sample size, in such settings even small-time savings can translate into increased availability of crews, improved response capacity, and reduced delays for subsequent patients – which in turn can literally be the difference between life and death or avoiding long-term disability. An observed increase in handover-to-clear time may reflect adjustments in documentation workflows, such as clinicians choosing to complete notes after patient handover, but might also reflect the fact that the technology was not integrated into existing systems. This highlights the importance of understanding behavioural adaptations and underlines the value of closer integration with electronic health record systems in future deployments.

Crucially, there were no compromises in safety or documentation quality across either setting. These findings reinforce those of earlier AVT evaluations in hospital environments, which similarly found increased clinical capacity without elevated risk.^19^ ^20^ The survey and interview findings indicated significant reductions in cognitive load and improved clinician experience and the potential for improved patient outcomes and experience, supporting findings in other clinical environments.^21–25^ Clinicians endorsed the introduction of AVT into routine practice, albeit with some further refinements required to templates and hardware, tailored training and a clear pathway to how improved efficiencies will be incorporated into clinician wellbeing approaches.^26^

### Strengths and weaknesses of the study

This evaluation represents the first study of its kind to assess the use of AVT across both remote telephone triage and on-scene emergency care within an NHS ambulance service. Building on previous AVT deployments in emergency departments and outpatient settings,^7^ ^8^ this pilot offers new, real-world evidence of how AI-assisted clinical documentation can improve frontline efficiency without compromising quality or safety. Critically, it is the first pilot that has sought to test AVT in the undifferentiated and multi-context setting of the ambulance service.

Application of this technology, given the breadth and complexity of both settings and clinical situations, represents a significant addition to knowledge and provides a foundation for further assessment and implementation of these technologies.

This evaluation was delivered within a compressed timeframe, limiting the scope of preparatory activity ahead of implementation. Interim technical workarounds, such as manual audio bridging in CHUB, may have influenced user experience and results. AVT was deployed as an emerging tool without full integration into existing systems. Importantly, full technical integration with existing clinical systems was not achieved during the evaluation period, which likely attenuated the potential productivity benefits of the technology. The deployment of AVT within these clinical environments represents a novel application of emerging technology. As such, optimal workflows, best practices, and effective integration strategies remain underdeveloped and are not yet fully established.

Participating clinicians were a small self-selecting cohort, introducing potential bias, although the direction of any impact is uncertain. Results may have been affected by seasonal demand fluctuations and varying patient acuity, especially when results are further segregated between conveyed/non-conveyed cases.

Sample sizes were not adjusted and may not fully reflect typical case mix or acuity so results should be interpreted in that context. Future evaluations should account for priority cases (e.g. patients requiring a hospital pre-alert) and ensure breath of inclusion of case-mix and types to assess system-wide applicability.

### Meaning of the study: implications for clinicians and policy-makers

This initial phase demonstrates clear proof of value of AVT in telephone triage and lays a strong foundation for further work in face-to-face ambulance applications. As the NHS continues to look for solutions that can release clinical time, reduce cognitive load, and improve workforce resilience, this evaluation provides compelling evidence that AVT, when implemented with effective clinical leadership and operational alignment, can play a meaningful role in shaping the future of urgent and emergency care delivery. In addition to releasing resources, supporting the diverse ambulance workforce and improving work-flows, this technology may unlock better clinical outcomes and timeliness of access to appropriate care within this highly complex and undifferentiated environment. This work offers timely and actionable insights into the role of AI in supporting ambulance services and reinforces the potential of AVT to help address capacity and access challenges facing the NHS.

### Future research

The data gathered will inform the design of a more extensive, fully integrated pilot. A larger-scale evaluation will be essential to explore longer-term impact, clinician adoption behaviours, and broader system-level benefits.

## Funding

The evaluation was supported by NHS England (London) Frontline Digitisation and Great Ormond Street Hospital DRIVE unit; the AVT platform (Tortus AI) was supplied free of charge. The GOSH DRIVE Unit receives infrastructure support from the NIHR GOSH Biomedical Research Centre and from the NIHR Healthtech Research Centre for Paediatrics and Child Health, which has made this work possible. London Ambulance Service NHS Trust technical and clinical project resourcing was provided for pilot implementation.

## Transparency statement

The lead author (Dr Shankar Sridharan) affirms that the manuscript is an honest, accurate, and transparent account of the study being reported; that no important aspects of the study have been omitted. There were no discrepancies from the study as originally planned.

## Declaration of Interests

Data collection, analysis and interpretation of findings were carried out by the LAS and GOSH teams independently of the technology partner. GOSH has entered a collaborative partnership with Tortus AI.

## Data Sharing

All quantitative findings reported are based on data from NHS service evaluations. Deidentified data are available upon reasonable request to the corresponding author.

## Data Availability

Deidentified data are available upon reasonable request to the corresponding author.

## Notes

### Author Declarations

The overall study was registered at GOSH as a clinical evaluation study and local approvement and study registration were additionally undertaken. NHS ethical approval was not required as per Andrew Pearson, the Clinical Audit Manager in the Quality Team at GOSH. Participating clinicians provided verbal consent; consent for surveys was assumed if a completed survey was submitted. Patients in the Hear and Treat pathway were informed that calls were being recorded and transcribed and that they could ask for transcription to be deactivated. Use of AVT in the face-to-face arm, where patients were present, was subject to verbal consent from patients and others on scene, including ambulance staff not in the trial. Some situations meant that clinicians were permitted to use discretion for AVT use depending on clinical circumstances, patient presentation (including mental capacity), or environmental factors.

