## Supplementary Table A and Figure A for "“What witchcraft is this?”: Paramedics report gains in productivity, well-being, and patient flow from piloting ambient voice technology in an NHS Ambulance Service"

**Supplementary Appendix**

### Table of Contents

**Box A: Stages of assessment of documentation quality in the Clinical Hub (CHUB) ....... 2**

**Figure A: 'What three words' at baseline (top) and AVT (bottom). .................................... 3**

**Box A:** Stages of assessment of documentation quality in the Clinical Hub (CHUB)

- All CHUB telephone consultations are routinely recorded, enabling detailed review of the assessment, clinician decision-making and resulting documentation.
- Each record is assessed and scored as a percentage.
- Additional audits for the pilot involved a trained manager listening back to the recording of the consultation, utilising the Clinical Guardian audit tool, allowing a consistent assessment and randomised allocation of audits to staff.
- Areas of assessment are set by the ambulance service and include review for accuracy and appropriateness of the clinical note documented on the record and the clinical decision making.

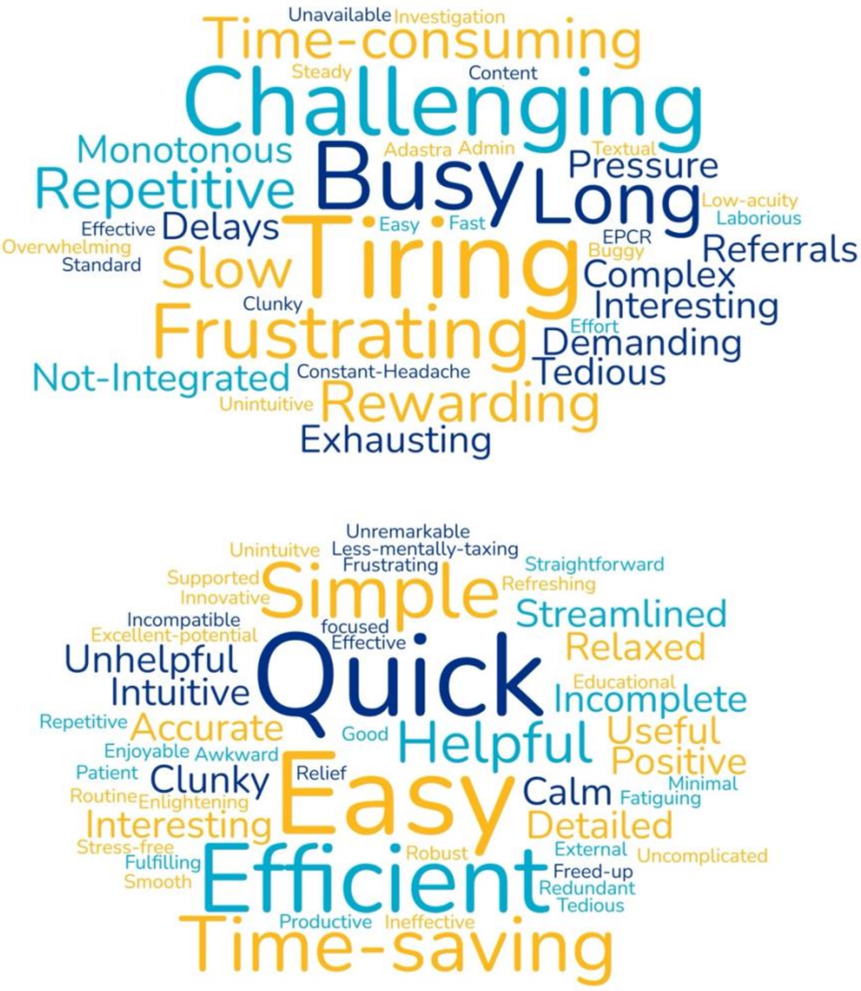


**Figure A**: 'What three words' at baseline (top) and AVT (bottom).

Footnote: Clinicians were asked to list the three words that described their experience of their most recent clinical shift at baseline and using AVT
